# Heterogeneity in statin responses explained by variation in the human gut microbiome

**DOI:** 10.1101/2021.12.02.21267193

**Authors:** Tomasz Wilmanski, Sergey A. Kornilov, Christian Diener, Mathew Conomos, Jennifer C. Lovejoy, Paola Sebastiani, Eric S. Orwoll, Leroy Hood, Nathan D. Price, Noa Rappaport, Andrew T. Magis, Sean M. Gibbons

## Abstract

Statins remain one of the most prescribed medications worldwide. While effective in decreasing atherosclerotic cardiovascular disease risk, statin use is associated with several side effects for a subset of patients, including disrupted metabolic control and increased risk of type II diabetes. We investigated the potential role of the gut microbiome in modifying patient response to statin therapy. In a cohort of >1840 individuals, we find that the hydrolyzed substrate for 3-hydroxy-3-methylglutarate-CoA (HMG-CoA) reductase, HMG, may serve as a reliable marker for statin on-target effects. Through exploring gut microbiome associations between blood-derived measures of statin effectiveness and metabolic health parameters among statin users and non-users, we find that heterogeneity in statin response is associated with variation in the gut microbiome. A *Bacteroides* rich, α-diversity depleted, microbiome composition corresponds to the strongest statin on-target response, but also greatest disruption to glucose homeostasis, indicating lower treatment doses and/or complementary therapies may be beneficial in those individuals. Our findings suggest a potential path towards personalizing statin treatment through gut microbiome monitoring.

## Introduction

Between 25% - 30% of older adults across the United States and Europe take statins regularly for the purpose of treating or preventing atherosclerotic cardiovascular disease (ACVD), making statins one of the most prescribed medications in the developed world ^1,2^. While statins have proven to be highly effective in decreasing ACVD-associated mortality, considerable heterogeneity exists in terms of efficacy (i.e., lowering low density lipoprotein (LDL) cholesterol) ^3^. Furthermore, statin use can give rise to a number of side effects in a subset of patients, including myopathy, disrupted glucose control, and increased risk of developing type II diabetes (T2D) ^4–8^. Several guidelines exist for which at-risk populations should be prescribed statins and at what intensity ^9^. However, despite considerable progress in identifying pharmacological ^10^ and genetic factors ^11^ contributing to heterogeneity in statin response, personalized approaches to statin therapy remain limited. Many times, treatment decisions are made through trial and error between the clinician and patient to obtain an optimal tolerable dose ^12^. Avoiding this trial-and-error phase through individualized analysis of genetic, physiological, and health parameters has the potential to improve drug tolerance, adherence, and long-term health benefits, as well as guide complementary therapies aimed at mitigating side effects.

Several studies have recently demonstrated a link between the gut microbiome and statin use ^13,14^. Similar to other prescription drugs, statins are widely metabolized by gut bacteria into secondary compounds ^15,16^. This indicates that the gut microbiome may impact statin bioavailability or potency to its host, contributing to the interindividual variability in LDL response seen among statin users ^17^. Additionally, biochemical modification of statins by gut bacteria could potentially contribute to side effects of the drug ^18^. Independent of statins, the gut microbiome has a well characterized role in contributing to host metabolic health through regulating insulin sensitivity, blood glucose, and inflammation, hence sharing considerable overlap with off-target effects of statin therapy ^19,20^.

Statin intake has also been implicated in shifting gut microbiome composition, where primarily obese individuals taking statins were less likely to be classified into a putative gut microbiome compositional state, or ‘enterotype’, defined by high relative abundance of *Bacteroides* and a depletion of short-chain fatty acid (SCFA) producing Firmicutes taxa ^21^. However, contradictory findings in animal models have also been reported, where a statin intervention decreased abundance of SCFA-producing taxa and, consequently, the gut ecosystem’s capacity to produce butyrate ^22^.

Given the numerous documented interactions between the gut microbiome and statins, and the established effect of the gut microbiome on metabolic health, we sought to explore the potential role of the gut microbiome in modifying the effect of statins on inhibiting their target enzyme 3-hydroxy-3-methylglutarate-CoA (HMG-CoA) reductase, as well as influencing the negative side effects of statins on metabolic health parameters. We analyzed data from over 1840 deeply-phenotyped individuals with extensive medication histories, clinical laboratory tests, plasma metabolomics, whole genome and stool 16S rRNA gene amplicon sequencing data. We found that heterogeneity in statin on-target effects and off-target metabolic disruption could be explained by variation in the composition of the gut microbiome. Overall, our results suggest that, with further study and refinement, the taxonomic composition of the gut microbiome may be used to inform personalized statin therapies.

## Results

### Cohort

The study population is presented in Fig. 1A and Table S1 (see also Methods). Briefly, a total of 1848 adults were included in the present analysis. Individuals in this cohort were self-enrolled in a now closed Scientific Wellness company (Arivale, Inc), had available plasma metabolomics and clinical laboratory data, and provided detailed information on prescription medication use. Of these 1848 Arivale participants, 244 were confidently identified as statin users, of which 97 provided detailed information on both dosage and type of statin prescribed.

**Fig. 1.**
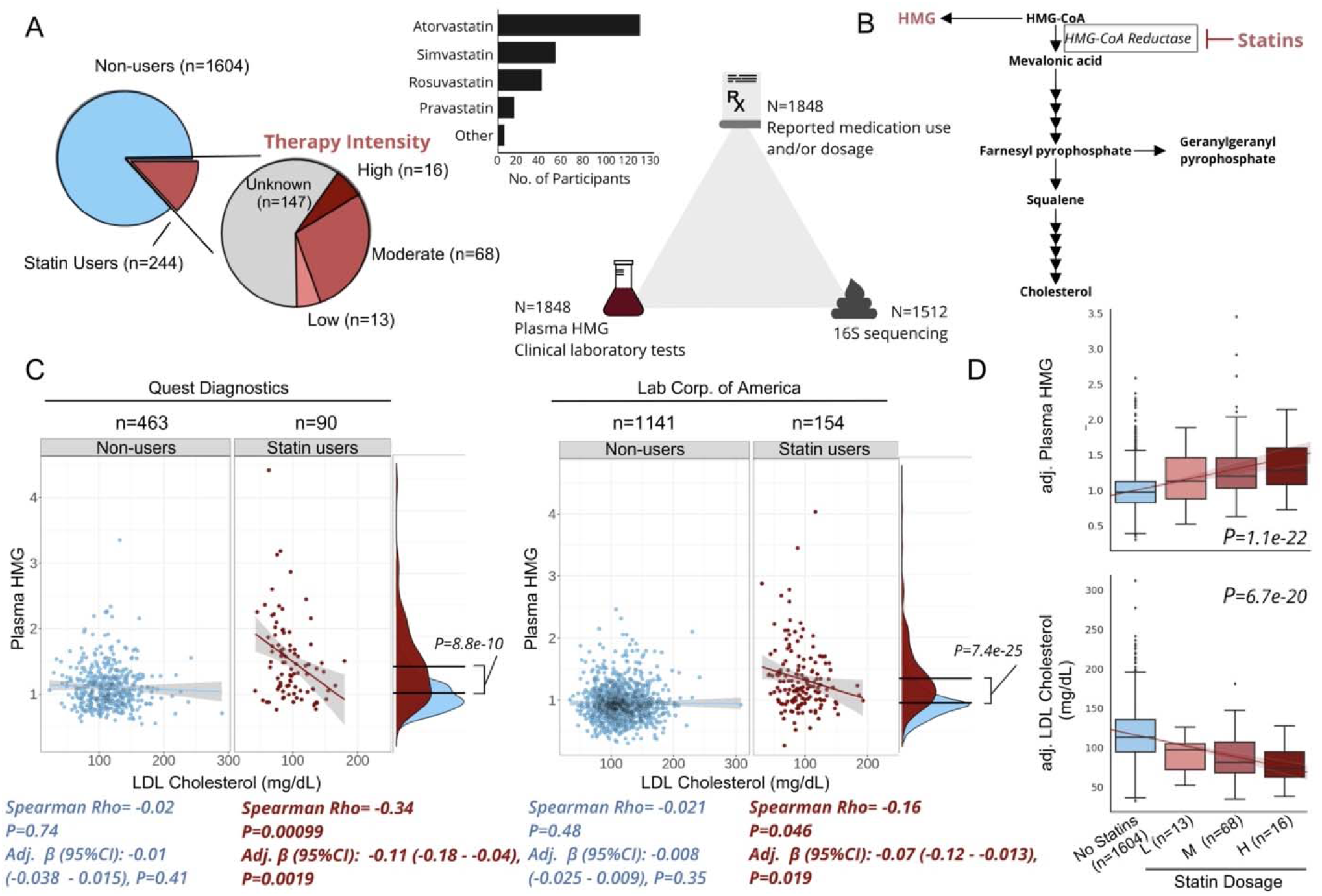
Plasma HMG correlates with statin use and statin LDL-response. **A)** Frequency of statin use, type of statin taken, and number of participants with available data for each ‘omics for each participant included in the present analysis. **B)** Diagram of *de novo* cholesterol synthesis pathway, with HMG and the rate-limiting enzyme inhibited by statins highlighted. **C)** Scatterplots of LDL-cholesterol and plasma HMG in statin non-users (blue) and users (red) separately, across two different clinical laboratory test vendors used in the cohort. The lines shown are the y∼x regression lines, and the shaded regions are 95% confidence intervals for the slope of each line. Below each scatter plot is the Spearman correlation coefficient and corresponding p-value. Adj. β(95%CI) corresponds to the β-coefficient for LDL cholesterol from GLMs predicting plasma HMG, adjusted for sex, age, and BMI. Also shown to the right of each scatter plot are kernel density plots for plasma HMG in statin users and non-users. The black lines indicate the mean of each group, and the p-value corresponds to the effect size of the difference between statin users and non-users from GLMs adjusted for the same covariates as above. **D)** Relationship between statin intensity therapy and plasma HMG as well LDL cholesterol levels for the subset of participants in the cohort who had available dosage intensity data (n=97). The lines shown are the y∼x regression lines, and the shaded regions are 95% confidence intervals for the slope of each line. P-value corresponds to the dose-response relationship between therapy intensity and either plasma HMG (top box plot) or LDL cholesterol (bottom box plot). Values on the y-axis are analyte levels adjusted for covariates (residuals). Box plots represent the interquartile range (25th to 75th percentile, IQR), with the middle line denoting the median; whiskers span 1.5 × IQR, points beyond this range are shown individually.

### Plasma HMG is a marker of statin use and on-target effects

The mechanism of action of statins is to inhibit the rate-limiting enzyme of *de novo* cholesterol synthesis, HMG-CoA reductase ^23^. Thus, we first sought to explore whether elevated plasma levels of the hydrolyzed substrate for this enzyme, HMG (measured in our broad untargeted metabolomics panel), could serve as a reliable marker of statin use (Fig. 1B). Plasma HMG levels were significantly higher in statin users than in non-users, consistent with our initial hypothesis and the drug’s well-established mechanism of action (Fig. 1C, generalized linear models (GLMs) adjusted for sex, age, and BMI, Quest Diagnostics β(95% confidence interval (CI)): 0.23 (0.16-0.31), *P=9*.*2e-10*), Lab Corp. of America (LCA) β(95% CI):0.28(0.23-0.34), *P=9*.*8e-25*). HMG levels further showed a negative correlation with blood LDL-cholesterol across two independent clinical lab vendors, but exclusively in statin users, indicating that plasma HMG may not only reflect statin use but also the extent to which statins inhibit their target enzyme (Fig. 1C, GLM adjusted for sex, age, and BMI, Quest β(95% CI): −0.12 (−0.19-0.05), *P=0*.*0019*), LCA β(95% CI):-0.07(−1.2 --0.01), *P=0*.*020*)).

The negative association between HMG and LDL-cholesterol, observed exclusively in statin users, indicates that this compound may serve as a proxy for statin efficacy. However, it is also possible HMG simply reflects patient adherence, where individuals who take the drug as prescribed have higher HMG and lower LDL-cholesterol than those who do not. To further evaluate the robustness of HMG as a marker for statin on-target effects, we explored its correspondence to variable doses of statins prescribed in a subset of statin users where this information was available (n=97). Different statins (atorvastatin, simvastatin, etc.) exhibit different potencies and are often prescribed at variable doses. To synchronize medical practices in terms of statin therapy, the American Heart Association (AHA) released guidelines for adjusting statin doses across all types of statins, which cluster into one of three intensity categories (low, moderate, and high) aimed at achieving desired decreases in LDL-cholesterol of <30%, 30-49%, ≥50%, respectively ^9^. Based on these AHA guidelines, a daily 40mg dose of Rosuvastatin would place a patient in the high intensity category, while the same dose of Fluvastatin would place a patient in the low intensity group. Hence, we re-classified participants into their respective therapy intensity groups based on the AHA guidelines (Fig. 1A) and evaluated the associations between therapy intensity, plasma HMG, and blood LDL-cholesterol levels. Therapy intensity showed a positive dose response relationship with HMG, independent of sex, age, and BMI (GLM adj. β(95% CI):0.15(0.12-0.17), *P=1*.*1e-22*)). Consistently, an inverse relationship was observed between therapy intensity and blood LDL-cholesterol (Fig. 1D, Ordinary Least Square (OLS) regression adjusted for sex, age, BMI and clinical lab vendor, β(95% CI):-15(−18 --12), P=*6*.*7e-20*).

Previous large-scale pharmacogenomic studies of statin users have identified several single nucleotide polymorphisms (SNPs) predisposing patients to variable responses to statin therapy. To evaluate if plasma HMG captures known genetic variability in statin response, we tested associations between HMG and 9 SNPs most strongly associated with statin-mediated decrease in LDL-cholesterol in previous studies ^11^, using GLMs with a statin-by-genetic variant interaction term while adjusting for sex, age, BMI and genetic ancestry (see Methods). Of the 9 SNPs tested, 2 SNPs in close linkage disequilibrium (rs445925 and rs7412 mapping to the APOC1 and APOE genes, respectively, *r* > 0.80 in Caucasians) showed significant associations with HMG, that were dependent on statin intake (i.e., the effect was only present in statin users, FDR<0.05), in the directions consistent with the previously described associations of the same variants with statin response (Fig. S1, Table S2). Interestingly, running the same analysis with LDL-cholesterol instead of plasma HMG as an outcome variable (both measured from the same blood draw) did not reveal the same statin-dependent interactions (Table S2). In the case of both rs445925 and rs7412, carrying at least one copy of the minor allele was associated with a decrease in LDL-cholesterol across statin users and non-users alike, hence providing no additional insight into statin-specific effects when measured cross-sectionally (Fig. S1). Together, our combined analyses of statin use, statin therapy intensity and genetic variants known to modify statin response indicate that HMG may provide additional insight into statin on-target effects, not captured by a snapshot measurement of LDL-cholesterol.

### Statin use is associated with subtle differences in the gut microbiome

Given the previously established associations between the gut microbiome and statin use, we next investigated whether statin intake is associated with changes in gut microbiome composition. Consistent with previous research, statin use showed a significant association with interindividual variability in gut microbiome composition, using the Bray-Curtis dissimilarity metric (PERMANOVA unadjusted model R^2^=0.0025, *P=0*.*00067*, model adjusted for microbiome vendor, sex, age, and BMI, R^2^=0.0021, *P=0*.*0017*) and Weighted UniFrac (unadjusted model R^2^=0.0017, P=0.031, model adjusted for the same covariates as the Bray-Curtis model, R^2^=0.0013, P=0.065) (Fig. 2A, Fig. S1). Statin intake was further associated with a modest decrease in one of the two α-diversity metrics calculated (OLS regression models predicting Shannon diversity adjusted for the same covariates as PERMANOVA, adj. β(95% CI):-0.095 (−0.16 --0.028), P=0.0051) (Fig. 2B). When looking at specific statin therapy intensity for a subset of participants where this information was available, there was no monotonic dose-response relationship with gut α-diversity, with only individuals receiving moderate intensity statin therapy demonstrating a significant decrease in measures of gut α-diversity relative to non-users (Fig. 2C, Fig. S1).

**Fig. 2.**
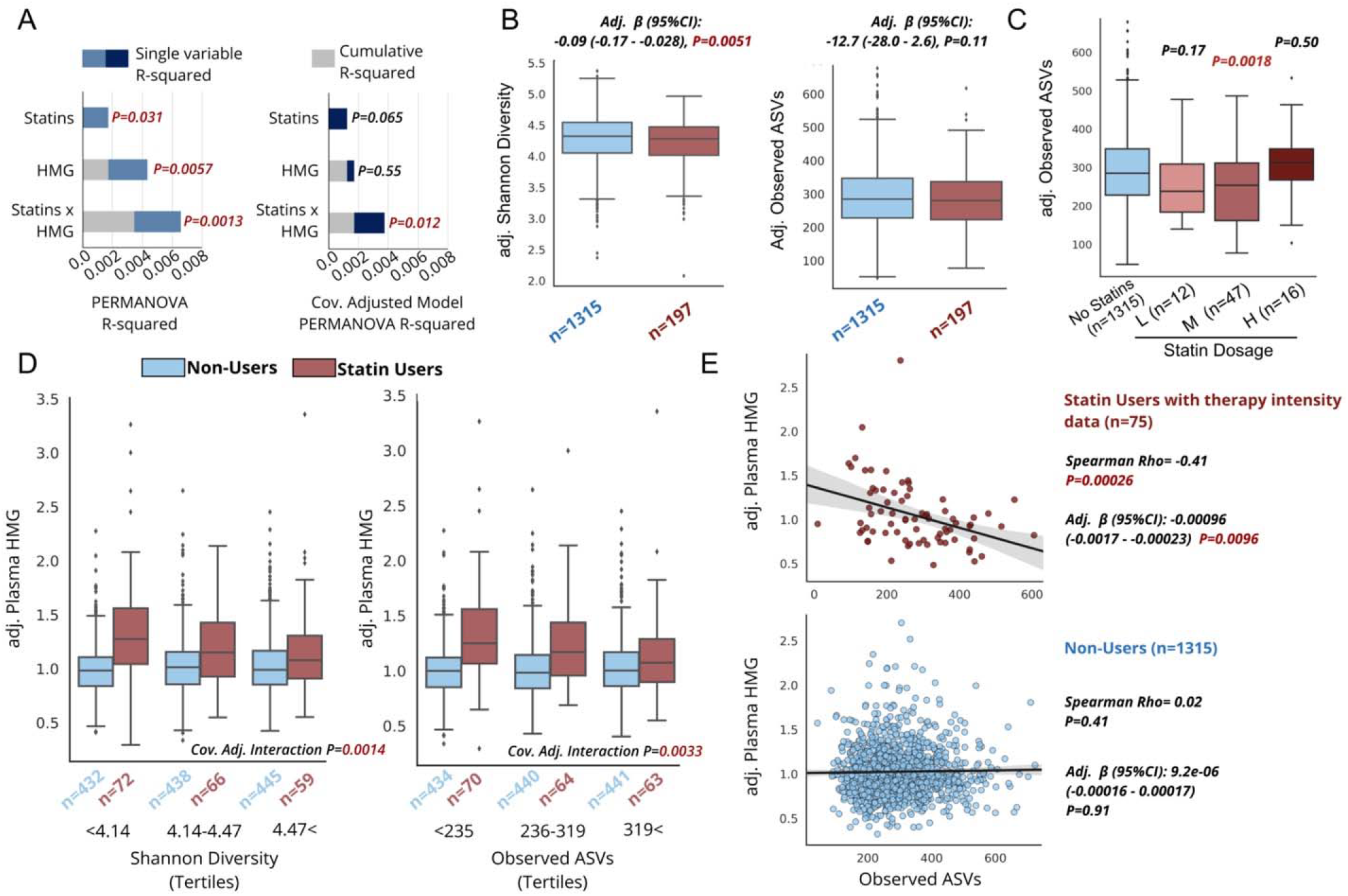
Gut microbiome composition is associated with markers of statin efficacy. **A)** Proportion of variance explained by statin use, plasma HMG levels, and a statin-by-HMG interaction term from unadjusted PERMANOVA models or models adjusted for sex, age, BMI, and microbiome vendor using the Weighted UniFrac genus-level dissimilarity matrix. Grey area corresponds to the cumulative R-squared of variables added to the model prior to the variable indicated on the x-axis, while the colored areas of the bars represent the additional variance explained by that variable. **B)** Measures of gut α-diversity in statin users compared to non-users. The β-coefficient, 95%CI and p-value shown is derived from OLS models predicting each of the α-diversity measures adjusted for microbiome vendor, sex, age, and BMI. Values on the y-axis are diversity measures adjusted for these covariates (residuals). **C)** Measures of Observed ASVs in statin users and non-users with known therapy intensity (low, moderate, high). P-values shown correspond to β-coefficients from OLS models predicting Observed ASVs comparing each intensity group to the no statin control group, adjusted for the same covariates as in B). **D)** Plasma HMG levels among statin users and non-users across tertiles of gut α-diversity. Interaction P corresponds to the statin-by-α-diversity interaction term p-value from GLM predicting plasma HMG adjusted for the same covariates as in B) and C). Values on the y-axis are diversity measures adjusted for these covariates (residuals). Box plots represent the interquartile range (25th to 75th percentile, IQR), with the middle line denoting the median; whiskers span 1.5 × IQR, points beyond this range are shown individually. **E)** Scatter plots of Observed ASVs (x-axis) and covariate adjusted plasma HMG levels (residuals) (y-axis) in statin users with known dosage therapy intensity as well as statin non-users. Levels were adjusted for the same covariates as in B), as well as dosage intensity.

### Gut microbiome α- and ß-diversity correlate with markers of statin efficacy

Next, we investigated whether gut microbiome beta-diversity may explain interindividual heterogeneity in response to statin therapy. Using HMG as a proxy for statin inhibition of its target enzyme, we modelled correspondence between statin on-target effects and interindividual variability in gut microbiome ß-diversity using PERMANOVA and including a statin-by-HMG interaction term. The interaction terms had permutation-based p-values of 0.0070 (R^2^=0.0017) and 0.0013 (R^2^=0.0032) for Bray-Curtis and Weighted UniFrac metrics, respectively, which remained significant after adjusting for microbiome vendor, BMI, sex, and age (Fig. 2A, Fig. S1). These results indicate that HMG correspondence to gut microbiome composition is dependent on statin intake, similar to the HMG-SNP associations reported earlier (Fig. S1). Very similar patterns were observed for gut α-diversity, where, once again, the association between our proxy for statin efficacy, HMG, and gut α-diversity was dependent on statin intake (Figure 2D). Plotting the association between gut α-diversity and HMG stratified by statin use revealed that, among statin users, higher α-diversity corresponded to lower plasma HMG levels, indicating decreased on-target effects of the drug in individuals with more diverse microbiomes (Fig. 2D). The negative association between HMG and α-diversity in statin users was also orthogonal to genetic variants predisposing individuals to variable statin responses. Running a stepwise forward regression model predicting HMG levels using the 9 SNPs previously associated with statin response explained an additional 3.2% of variance in HMG, on top of age (i.e. the base model). Including observed ASVs as a measure of gut diversity in the model in addition to age and the chosen SNPs increased the percent variance explained by an additional 3.9% (complete model R^2^=0.185).

To further exclude the possibility that individuals with higher α-diversity are generally healthier and simply prescribed less potent statin therapies to begin with, thus leading to lower levels of HMG, we further adjusted our models for dosage intensity in the subset of participants with microbiome data where this information was available (n=75). In this smaller group of individuals, associations between gut α-diversity and HMG were not impacted by correcting for statin intensity (Fig. 2E & Fig. S2). Similar results were observed when investigating statin dependent associations between LDL-cholesterol and gut α-diversity, although to a weaker extent (OLS models predicting LDL-cholesterol adjusted for clinical lab and microbiome vendors, sex, age, and BMI, statin-by-Shannon diversity interaction term β(95% CI):12.2(2.5-22.0), *P=0*.*014*, statin-by-Observed Amplicon Sequence Variants (ASVs) interaction term β(95% CI):0.042(0.00086-0.084), *P=0*.*044*, Fig. S2). A weaker interaction effect with LDL cholesterol is to be expected, given the cross-sectional nature of our study and our inability to capture the percent decrease in LDL-cholesterol from baseline following the initiation of statin treatment, one of the most common and direct measures of statin effectiveness ^3,17^.

As another measure of gut microbiome correspondence to statin response, we tested the association between measures of gut α-diversity and the likelihood of having reached pre-defined target LDL-cholesterol levels for statin users (<70mg/dL and <100mg/dL). These are clinically relevant targets, as clinicians are recommended to adjust dosage and type of statin prescribed to reach these levels of LDL-cholesterol depending on the presence of specific ASCVD risk factors in their patients ^24^. Both Shannon diversity and Observed ASVs showed negative associations with likelihood of having reached target LDL-levels among statin users (Multivariable logistic regression adjusted for clinical lab vendor, sex, age, BMI, and T2D status [a common criteria, in combination with one or more CVD risk factors, where more aggressive LDL-lowering therapy is pursued]): Odds Ratios (OR) ranging from 0.60-0.69, Table 1). Together, these results indicate that gut microbiome composition can explain a significant proportion of variability in statin on-target effects in a generally healthy community-dwelling population.

**Table 1.** Gut microbiome measures correlate with having reached LDL-cholesterol target levels among statin users. Odds Ratios (OR) for each gut microbiome measure from logistic regression models predicting having achieved either <100 mg/dL or <70 mg/dL target LDL-cholesterol level among statin users. The *Bac*.*2* enterotype was compared against all other enterotypes. Measures of α-diversity were scaled and centered prior to analysis for easier comparison of effect sizes. Models were adjusted for clinical laboratory and microbiome vendors, age, sex, and BMI. Further adjustment for T2D status was done in participants where this information was available (n=174). Significant OR (P<0.05) are highlighted in red.

|  | <100 mg/dL (n cases=132, N total=197) |  | <70 mg/dL (n cases=44, N total=197) |  |
| --- | --- | --- | --- | --- |
|  | Cov. adj.<br>OR(95%CI) | Cov. & T2D adj.<br>OR(95%CI) | Cov. adj.<br>OR(95%CI) | Cov. & T2D adj.<br>OR(95%CI) |
| Shannon diversity | 0.69 (0.49-0.97) | 0.72 (0.50-1.03) | 0.67 (0.48-0.95) | 0.60 (0.41-0.87) |
| Observed ASVs | 0.67 (0.47-0.95) | 0.67 (0.45-0.98) | 0.66 (0.45-0.95) | 0.62 (0.40-0.96) |
| Bac.2 enterotype | 2.19 (1.04-4.60) | 2.11 (0.95-4.66) | 3.61 (1.68-7.77) | 4.33 (1.83-10.25) |

### Statin-associated changes in on- and off-target effects are dependent on microbiome compositional states

Prior work on the gut microbiome and statins has relied on clustering individuals into microbiome-based compositional states called ‘enterotypes’ ^25,26^. A recent study revealed that statin intake among obese individuals was associated with lower prevalence of the *Bacteroides* 2 (*Bac*.*2*) enterotype, which is generally considered to be less healthy than other broad enterotype groupings common to cohorts in the United States and Europe ^21^. To evaluate the extent to which these coarse ecological groupings might help explain interindividual variation in statin on- and off-target effects, we stratified our cohort into enterotypes. Using a previously established method for enterotype identification, Dirichlet multinomial mixture (DMM) modeling ^27^, the participants in the Arivale cohort separated optimally into four groups, according to the Bayesian Information Criterion (BIC), consistent with some, but not all, previous human gut microbiome studies (*Bacteroides* 1 (*Bac*.*1*), *Bac*.*2, Ruminococcaceae* (*Rum*.), and *Prevotella* (*Prev*.) clusters) ^21,26–28^ (Fig. 3A, Fig. S2). The four enterotypes identified showed very similar characteristics to those described previously in European cohorts, with two *Bacteroides*-dominated enterotypes (*Bac*.*1* and *Bac*.*2*), with the *Bac*.*2* enterotype being further characterized by decreased α-diversity and a depletion of SCFA-producing commensals like *Faecalibacterium* and *Subdoligranulum* (Fig. 3B, Fig. S2). The *Rum*. enterotype was enriched for taxa primarily from the Firmicutes phylum, as well as *Akkermansia* (Fig. S2, Data S1), consistent with previous findings ^25^. The *Prev*. enterotype was the smallest in size and characterized by high relative abundance of the *Prevotella* genus (Fig. 3D, Data S1).

**Fig. 3:**
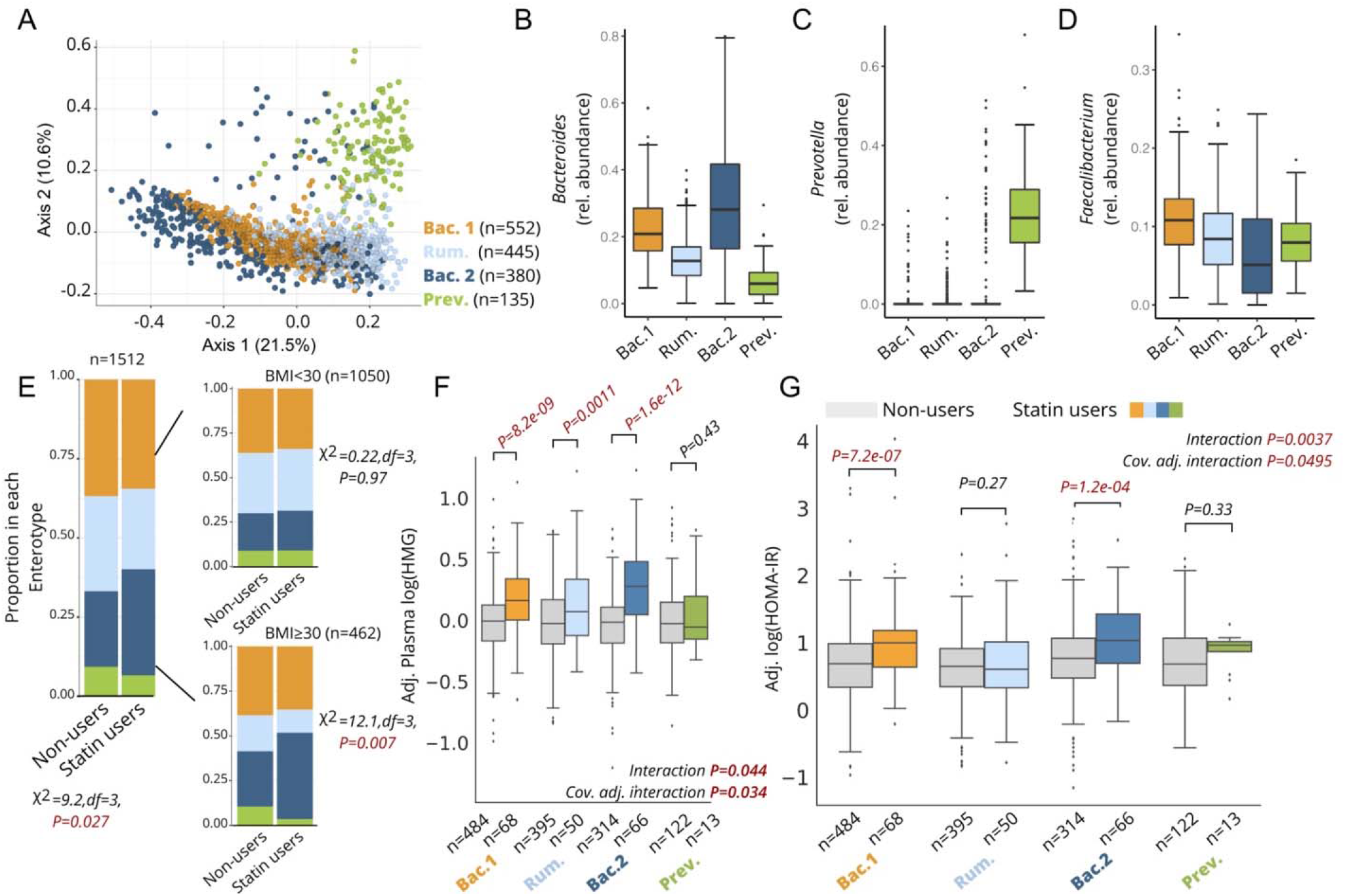
Statin associations with markers of efficacy and metabolic side effects are microbiome enterotype dependent. **A)** Principal Coordinate Analysis (PCoA) plot of the genus-level Bray-Curtis Dissimilarity matrix color-coded by enterotypes. **B-D)** Relative abundance of *Bacteroides* (B), *Prevotella* (C), and *Faecalibacterium* (D) genera across the four enterotypes identified in the cohort. **E)** Proportion of each enterotype in statin users and non-users across the whole cohort (left) and stratified by obesity status (right). **F)** Plasma HMG levels among statin users and non-users stratified by enterotype. Interaction P corresponds to the statin-by-enterotype interaction term p-value from unadjusted ANOVA models, while the cov. Adj. interaction P corresponds to the statin-by-enterotype interaction term p-value from covariate adjusted ANCOVA models. Plasma HMG levels shown on the y-axis are values adjusted for the same covariates (residuals). **G)** HOMA-IR measures among statin users and non-users stratified by enterotype. Interaction P corresponds to the statin-by-enterotype interaction term p-value from unadjusted ANOVA models, while the cov. Adj. interaction P corresponds to the statin-by-enterotype interaction term p-value from ANCOVA models adjusted for covariates. P-values above the box plots in F) and G) correspond to tests of significance between statin non-users and statin users within each enterotype using two-samples t-test. Differences with Bonferroni corrected *P<0*.*05* were considered statistically significant and are highlighted in red. Box plots represent the interquartile range (25th to 75th percentile, IQR), with the middle line denoting the median; whiskers span 1.5 × IQR, points beyond this range are shown individually.

We first attempted to replicate previous findings ^21^ documenting an observed lower prevalence of the *Bac*.*2* enterotype in obese individuals taking statins. Consistent with previous results, obesity itself was associated with a higher likelihood of being assigned to the *Bac*.*2* enterotype (Multivariable logistic regression adjusted for microbiome vendor, sex, and age, OR(95%CI): 1.8 (1.4-2.3), *P=5*.*0e-5*). However, contrary to the original findings, we actually observed a higher prevalence of the *Bac*.*2* enterotype among statin users compared to non-users, particularly among obese individuals (Fig. 3E). This association among obese individuals was further confirmed using multivariable logistic regression adjusting for sex, age, and microbiome vendor (OR (95%CI): 2.1 (1.2-3.7), *P=0*.*013*, n=462).

We next set out to explore whether an individual’s enterotype was associated with their response to statin therapy. Focusing on statin on-target effects, we observed a significant enterotype-by-statin interaction when modeling blood HMG levels (*P=0*.*044*, unadjusted analysis of variance (ANOVA), *P=0*.*034*, analysis of covariance (ANCOVA) adjusted for microbiome vendor, clinical lab vendor, sex, age, and BMI). Stratifying the cohort by enterotypes and comparing statin users to non-users revealed statin use within the *Bac*.*2* enterotype correlated with the highest HMG levels (37% mean increase relative to non-users), followed by the *Bac*.*1* (24%) and *Rum*. enterotypes (18%). Interestingly, individuals with a *Prev*. enterotype showed no significant difference in HMG between statin users and non-users, although our sample size for this enterotype was small and thus this result needs to be interpreted with caution (Fig. 3F). Similar results were obtained when evaluating statin-by-enterotype interaction effects on LDL-cholesterol levels (P=0.021, unadjusted ANOVA, P=0.0032, ANCOVA adjusted for same covariates as HMG models), with statin users with a *Bac*.*2* enterotype demonstrating lowest LDL-cholesterol levels (−33%) relative to non-users within the same enterotype (Fig. S3). Statin users who were assigned the *Bac*.*2* enterotype were also two to four-times more likely to have reached common LDL-cholesterol target levels for statin-users at higher risk for ASCVD (Table 1). Collectively, these results suggest that microbiome enterotypes may reflect the extent to which statins inhibit HMG-CoA reductase and reduce LDL-cholesterol levels across individuals.

Statin use has previously been associated with disrupted glucose control and increased risk of developing T2D in a subset of patients ^5,7,29^. Given the known role of the gut microbiome in contributing to metabolic homeostasis, and the variable metabolic profiles previously observed across different microbiome enterotypes ^21,30^, we investigated whether enterotypes may modify the association between statin use and markers of insulin resistance. Focusing initially on Homeostatic Model Assessment for Insulin Resistance (HOMA-IR) ^31,32^, we tested for an enterotype-by-statin interaction effect while adjusting for microbiome vendor, clinical lab vendor, sex, age, BMI, LDL-cholesterol, and plasma HMG using ANCOVA. Individuals showed variable responses to statin therapy based on their microbiome enterotype, with *Bac*.*2* individuals on statins demonstrating the highest levels of HOMA-IR relative to non-statin users, while *Rum*. individuals showed no significant difference in HOMA-IR between statin users and non-users (ANOVA unadjusted interaction term P=0.0037, ANCOVA covariate adjusted Interaction term P=0.0495, Fig. 3G, Table 2). It is worth noting that in the subset of participants where dosage intensity information was available, all three intensities (low, moderate, high) were associated with a comparable increase in HOMA-IR, suggesting that differences in therapy intensity are likely not the main driver behind the observed statin-enterotype interaction (Fig. S2).

**Table 2.** Gut microbiome enterotypes modify the association between statin use and markers of glucose homeostasis. Percent median increase in the first four columns corresponds to the percent difference in each marker between statin users and non-users within each enterotype. P-values in these columns correspond to t-tests comparing covariate adjusted values between statin users and non-users. Values shown are raw p-values, and those that remained significant after correcting for type-1-error (Bonferroni *P<0*.*05*) are highlighted in red. The last three columns in the table show the F- and p-values for the statin-by-enterotype interaction term from ANOVA (unadjusted) and ANCOVA (covariate adjusted) models predicting each of the specified markers of glucose homeostasis. Covariate adjusted models were adjusted for microbiome vendor, clinical lab vendor, sex, age, BMI, LDL cholesterol and plasma HMG. Last column corresponds to models adjusted for the same covariates as well as T2D status (yes/no, N=1691, T2D n=64). P-values<0.05 are colored in red. Abbreviations: HOMA-IR: Homeostatic Model Assessment for Insulin Resistance; HbA1c: Glycated Hemoglobin A1c.

| Measure | Percent median increase in each measure and P-value<br>Between statin-users and non-users for each enterotype |  |  |  | F-value and corresponding P-value for statin*enterotype interaction term predicting each measure |  |  |
| --- | --- | --- | --- | --- | --- | --- | --- |
|  | *Bac.1* | *Rum.* | *Bac.2* | *Prev.* | Unadjusted model<br>N=1848 | Covariate adj. model N=1848 | Covariate and diabetes adj. model N=1691 |
| HOMA-IR | 73%,<br>*P=7.2e-07* | 21%<br>*P=0.27* | 99%<br>*P=1.2e-04* | 29%<br>*P=0.33* | F=4.5,<br>*P=0.0037* | F=2.6,<br>*P=0.0495* | F=2.6,<br>*P=0.049* |
| Insulin | 63%<br>*P=5.6e-06* | 19%<br>*P=0.17* | 89%<br>*P=9.1e-04* | 22%<br>*P=0.25* | F=3.0,<br>*P=0.032* | F=1.4,<br>*P=0.23* | F=1.5,<br>*P=0.22* |
| Glucose | 6.6%<br>*P=9.7e-04* | 4.5%<br>*P=0.51* | 9.3%<br>*P=8.1e-04* | 7.6%<br>*P=0.84* | F=6.4,<br>*P=0.00025* | F=4.4,<br>*P=0.0041* | F=3.9,<br>*P=0.0092* |
| HbA1c | 5.6%<br>*P=2.0e-03* | 1.9%<br>*P=0.16* | 7.3%<br>*P=1.2e-04* | 1.8%<br>*P=0.57* | F=8.1,<br>*P=2.3E-05* | F=6.3,<br>*P=0.00030* | F=3.4,<br>*P=0.017* |

We next expanded our analysis into additional markers of metabolic health, including fasting insulin and blood glucose, as well as glycated hemoglobin A1c. There was a significant enterotype-by-statin interaction across all tested metabolic parameters, which remained significant after adjusting for covariates across all markers other than insulin (Table 1, Fig. S3). As individuals with T2D are often recommended to take statins, we further adjusted all models for T2D status in participants where this information was available (N=1691, T2D n=66), which did not change the significance of enterotype-by-statin interaction effects observed (Table 2). Collectively, these results suggest that gut microbiome composition may modify how statins influence off-target physiology, particularly glucose homeostasis.

## Discussion

There is considerable heterogeneity in response to statin therapy among individuals, both in terms of on-target effects (lowering LDL-cholesterol) and likelihood of experiencing unwanted side-effects ^3,7,29^. Herein, we report that variation in gut microbiome taxonomic composition can explain interindividual variability in statin responses. The main findings of our analyses are as follows: 1) HMG measured in plasma is a robust marker of both statin use and statin on-target effects, which also reflects known genetic variability in statin responses; 2) Gut α-diversity negatively correlates with HMG exclusively in statin users, independent of dose intensity and genetic predisposition, indicating a more diverse microbiome may interfere with statin on-target effects; 3) Enterotype analysis further confirms similar patterns of microbiome modification of statin response, with the *Bacteroides* dominant, α-diversity-depleted *Bac*.*2* enterotype showing the highest plasma HMG and lowest LDL-cholesterol levels among statin users; and 4) Of the four enterotypes identified, individuals with the *Bac*.*2* followed by *Bac*.*1* enterotypes experience greatest disruption to glucose control when on statins, while the Firmicutes rich *Rum*. enterotype appears most protective, indicating variable risk of statin-mediated metabolic side effects based on gut microbiome composition. Collectively, our findings indicate that the gut microbiome influences statin actions. With further refinement, knowledge of these effects may inform statin therapy guidelines and help personalize ASCVD treatment.

To the best of our knowledge, measuring HMG in large observational studies for the purpose of exploring statin-mediated effects has not been previously proposed. The conversion of HMG-CoA to HMG is dependent on the hydrolysis of the thioester bond linking HMG to its Coenzyme-A moiety, which has been previously shown to be facilitated by at least one known thioesterase (peroxisomal acyl-CoA thioesterase 2) ^33^. Relatively little is known about the accumulation of HMG with statin therapy and the pathways involved, which warrants further research. Nevertheless, there are several advantages for including HMG along with LDL-cholesterol measurements when evaluating statin effects. For one, given the limitations of a cross-sectional study design like ours, HMG may provide more time-invariant insight into statin efficacy, as opposed to LDL-cholesterol, which would require knowledge of pre-statin cholesterol levels to calculate the percent decrease in LDL over time ^3^. This seemed to be the case in our genetics analysis, where cross-sectional measurements of plasma HMG were able to capture previously reported genetic variability in statin response while LDL-cholesterol measurements from the same blood draw were less sensitive. In addition, plasma HMG could prove useful when evaluating statin off-target effects on metabolic health parameters, where statistical models could be adjusted for HMG to account for variability in statin on-target effects, as was done in our analysis exploring markers of insulin resistance.

An intriguing finding in the present analysis was an absence of statin-associated metabolic disruption in individuals with a *Rum*. enterotype (Fig. 3G, Fig. S3). Statin use in this group was still associated with increased plasma HMG and decreased LDL-cholesterol levels (Fig. 3F, Fig. S3), indicating that patients with this microbiome composition type may benefit from statin therapy without an increased risk of unwanted metabolic complications. There are several possible explanations for this observation. For example, the *Rum*. enterotype is enriched in the genus *Akkermansia*, as well as several butyrate-producing taxa, which are known to positively impact host metabolism through multiple mechanisms (Table S2, Fig. S2) ^25,34^, potentially serving as a buffer against statin off-target effects on glucose homeostasis. In addition, statins and other prescription drugs have been previously shown to be most readily metabolized by species within the *Bacteroides* genus, of which the *Rum*. enterotype is most depleted. The lower degree of drug metabolism by Firmicutes taxa comprising the *Rum*. enterotype may therefore be potentially protective from statin off-target effects. Consistently, both *Bacteroides* rich *Bac*.*1* and *Bac*.*2* enterotypes showed highest levels of markers of insulin resistance with statin use.

Statin use in individuals with the *Bac*.*2* enterotype was associated with the strongest on-target effects (i.e., high plasma HMG and low LDL-cholesterol levels) but also greatest metabolic disruption among all four enterotypes (Fig. 3F-G, Fig. S3). This is consistent with previous observational studies that have identified an association between the magnitude of decrease in LDL-cholesterol with statin use and risk of developing T2D (i.e., the greater the percent decrease in LDL-cholesterol with statin therapy, the higher the risk of new onset T2D) ^6,35^. One possible mechanism behind the reported association is the previously mentioned ability of *Bacteroides* species to metabolize prescription drugs, including statins ^15^. *Bacteroides* dominance within both the *Bac*.*1* and *Bac*.*2* enterotypes may modify drug activity, impacting both potency and potential side effects. Paired with depletion of several major butyrate-producing taxa within the *Bac*.*2* enterotype (Fig. 3D, Fig. S1, Data S1), this bacterial composition may put patients at particularly high risk of metabolic complications. If this were indeed the case, individuals with a *Bac*.2 enterotype could benefit most from lower intensity therapy, which might still achieve the desired percent decrease in LDL-cholesterol while mitigating potential metabolic disruptions. Complementary pro- or prebiotic interventions could also be potentially pursued in these individuals. However, further experimental work is needed to fully elucidate the microbiome-statin interactions that may be driving the reported associations.

While our present investigation identified very similar enterotype structure to a previous study on statin use and the gut microbiome by Veiera-Silva *et al*. ^21^, our analysis also showed conflicting results in terms of prevalence of the putatively dysbiotic *Bac*.*2* enterotype among obese statin users (Fig. 3E). One possible explanation for this discrepancy is that in the original study individuals were primarily prescribed Simvastatin (48% of statin users), which is a lower intensity HMG-CoA reductase inhibitor than the most commonly prescribed Atorvastatin in our cohort (53% of all statin users). The different statin types may have a different impact on the gut microbiome. Alternatively, the studied population of obese statin users in the original study by Veiera-Silva *et al*. may have been healthier and hence prescribed lower intensity therapies than the cohort presented in this study, resulting in these variable findings. Finally, additional confounding variables may be responsible for the observed shifts in enterotype proportions among obese statin users across both studies. It’s worth noting that, while the prior study ^21^ focused on how statins might influence the composition of the microbiome, our study focused on how the composition of the microbiome impacts the on- and off-target effects of statins in the host. Our analyses indicate that statins have a detectable, but very weak effect on the composition of the gut microbiome, while the gut microbiome appears to have a more sizable impact on host responses to statin treatment.

Growing evidence suggests a bidirectional interaction between prescription medication use and the gut microbiome, which may inform drug treatment for hundreds of millions of people worldwide. Here we present a proof-of-concept study on how gut microbiome composition may be used to stratify patients to inform statin therapy. As our understanding of microbe-drug interactions deepens, gut microbiome modification and monitoring hold promise for informing pharmacological treatment optimization.

## Online Methods

### Cohort

The Arivale cohort consists of adults (18+ years old) who self-enrolled in a now closed lifestyle intervention program (Arivale, Inc. 2015-2019) ^36–39^. The lifestyle intervention was designed to improve a number of key outcomes based on longitudinal profiling of clinical biomarkers and individualized coaching. For the present study, only individuals who filled out medication questionnaires, and/or reported their prescription medication information directly to their coach during a 1-on-1 session, were included. Participants further had to have available fasting plasma metabolomics and clinical laboratory test data (N=1848). Only baseline measurements and corresponding medication doses at the start of the program were considered, i.e. before any lifestyle interventions were recommended. Of the 1848 participants originally included, after excluding individuals who reported taking antibiotics in that last 3 months, 1512 had available stool 16S rRNA gene sequencing data. Similar to the larger Arivale population, the majority of participants of this study were residents of Washington and California when in the program. Although the cohort tends to be leaner than the general U.S. population (prevalence of obesity is 31% relative to the national prevalence of 42%^40^), it is representative of the populations in the states where the majority of the participants were located. The cohort is further predominantly female (63%) and is skewed towards Caucasians (81%). Additional demographic information on the cohort is provided in Table S1.

### Microbiome Analysis

Stool samples in the Arivale cohort were collected using kits developed by two microbiome vendors (DNAGenotek or Second Genome), and processed as described previously ^37,41^. Briefly, stool sample collection kits with proprietary chemical DNA stabilizers to maintain DNA integrity at ambient temperatures were shipped directly to participants’ homes and then shipped back to the vendors. Gut microbiome sequencing data in the form of FASTQ files were then obtained from the vendors on the basis of either the 300-bp paired-end MiSeq profiling of the 16S V3 + V4 region (DNAgenotek) or 250-bp paired-end MiSeq profiling of the 16S V4 region (Second Genome). Downstream analysis was performed using a denoise workflow from mbtools (https://github.com/gibbons-lab/mbtools) that wraps functions from DADA2. DADA2 ^42^ error models were first trained separately for each sequencing run and subsequently used to obtain amplicon sequence variants (ASVs) for each sample. Next, chimera removal was performed using the *de novo* DADA2 algorithm, which removed ∼17% of all reads. Taxonomy assignment was performed using the RDP classifier with the SILVA database (version 132). In summary, 99% of the reads could be classified to the family level, 89% to the genus level and 32% to the species level. Sequence variants were aligned to each other using DECIPHER ^43^ and multiple sequence alignment was trimmed by removing each position that consisted of more than 50% gaps. The resulting core alignment was then used to reconstruct a phylogenetic tree using FastTree ^44^. Downstream gut microbiome analysis was conducted using the *Phyloseq* Package in R (v 3.6) ^45^. Gut microbiome samples were first rarefied to an even sampling depth of 25596 reads, corresponding to the minimum number of reads per sample in the dataset. Bray-Curtis ^46^ and Weighted UniFrac ^47^ dissimilarity matrices were calculated at the genus-level using the *Phyloseq* package. Alpha-diversity measures were calculated at the ASV-level using the *Phyloseq* Package. Enterotype analysis was performed using Dirichlet Multinomial Mixture (DMM) modeling on the rarefied genus-level count data, as previously described ^27^. For selecting the optimal number of DMM groups in our cohort (i.e. enterotypes), we used the Bayesian information criterion (BIC) (Fig.S2).

However, BIC as a model penalization metric is not without limitations and tends to err on the side of underfitting (i.e., estimating a smaller number of clusters). The Laplace approximation for model penalization ^27^, on the other hand, did not identify an optimal number of clusters in this particular dataset (out to a maximal number of eight clusters tested), indicating limited statistical evidence for a small number of coarse-grained compositional states within our cohort (Fig. S2). Nevertheless, the main enterotype groupings tend to be relatively consistent from study-to-study in large U.S. and European populations, even if the statistical evidence for such states is somewhat limited ^26^. Given that the four BIC-identified enterotypes in our cohort show strikingly similar taxonomic signatures to those identified in prior work on statins ^21^, we moved forward with an analysis of these compositional states and how they relate to statin response.

### Clinical Laboratory Tests

Blood draws for all assays were performed by trained phlebotomists at LabCorp (n=1309) or Quest (n=553) service centers, and assaying was performed in CLIA-certified laboratory facilities by the vendors. Blood samples for clinical laboratory tests were obtained at the same time as the metabolomics blood draw, and only the baseline sample prior to any lifestyle coaching intervention was considered. Prior to the blood draw, Arivale participants were advised to avoid alcohol, vigorous exercise, aspartame, and monosodium glutamate for 24 hours, and to begin fasting 12 hours in advance.

### Plasma Metabolomics

Plasma HMG was measured as part of the metabolomics data generated by Metabolon, Inc., on the same blood draws as clinical laboratory tests. Raw metabolomics data was normalized as described previously ^36,37^. Values were median scaled within each batch, such that the median value for each metabolite was 1. To adjust for possible batch effects, further normalization across batches was performed by dividing the median-scaled value of each metabolite by the corresponding average value for the same metabolite in technical control samples processed in the same batch. The same technical control samples were used to ensure the comparability of abundance estimates obtained across batches.

### Genetics Analysis

Participant DNA was extracted from whole blood and, following quality control and purification, as needed, underwent 150 PE whole genome sequencing (WGS) using Illumina’s HiSeq X at 30x coverage as described previously ^48^. Variant calling was performed by the vendor using the pipeline that follows GATK’s Best Practices, using Haplotype Caller and hg19 build as the reference genome. A total of 1747 participants (∼94% of the present cohort) had available WGS data and were used in the present analysis. Following extensive quality control and assurance, genetic ancestry was calculated as principal components (PCs) using a set of ∼100,000 ancestry-informative SNP markers as described previously ^49^. SNPs chosen for testing associations with HMG were based on prior studies investigating genetic predisposition to statin efficacy defined as percent decrease in LDL-cholesterol from baseline, and included the following variants: rs10455872, rs2199936, rs2900478, rs4420638, rs445925, rs5908, rs646776, rs7412, and rs8014194 ^11^. To model the association between SNPs and HMG in statin users, individuals homozygous and heterozygous for the minor allele were grouped together. Statistical analysis was performed on each SNP individually using GLM with a Gamma distribution and a log-link function within the statsmodels module in Python, with HMG as the dependent variable and a statin-by-SNP interaction term. The interaction term tests for a significant association between HMG and statin use, that is modified by the SNP of interest (i.e. the effect of statins on HMG are variable based on the genetic variant). Models were further adjusted for sex, age, BMI and the first 7 ancestry PCs. Ordinary Least Square (OLS) regression models with the same covariates and interaction term were also run with LDL-cholesterol as the dependent variable. Type-1 error was controlled using the Benjamini-Hochberg method (FDR*<0*.*05*).

### Statistical Analysis

Depending on the statistical approach, analysis was conducted using either R (v 3.6) or Python (v 3.7). Of the 1848 participants included in our study, 73 had missing data on sex and age, 66 on BMI, and 81 on HMG. These missing values were imputed using plasma metabolomics data and the K nearest neighbor algorithm implemented through the sklearn.impute module in Python. The associations of plasma HMG levels with LDL-cholesterol, statin intensity, and measures of gut alpha-diversity were all tested using Generalized Linear Models (GLM) with a Gamma distribution and a log-link function within the statsmodels module in Python, with HMG as the dependent variable. OLS regression (Python) was used whenever LDL-cholesterol or measures of gut alpha-diversity were the dependent variables. Testing for associations between variables and interindividual variability in gut microbiome composition was conducted using permutational multivariate analysis of variance (PERMANOVA) through the *Adonis* package in R using both the genus-level Bray-Curtis and Weighted UniFrac dissimilarity matrices. The number of permutations to obtain P-values was set to 3000.

For assessing dose-response relationships between HMG/LDL-cholesterol and dosage intensity (Fig. 1D), dosage was recoded into an ordinal variable (0(none/no statins), 1(low), 2(moderate), 3(high)), and the significance of the β-coefficient for that variable from covariate adjusted models predicting either HMG(GLM adjusted for sex, age, and BMI) or LDL-cholesterol (OLS adjusted for sex, age, BMI, and clinical lab vendor) was reported. Wherever associations were visualized using box plots or scatter plots, the residuals (values adjusted for covariates from either GLM or OLS models) were plotted instead of the original values. For comparing the differences in prevalence of the four enterotypes among statin users and non-users, the **χ**^**2**^ test was performed using the chisq.test function in R. When evaluating the association between obesity and *Bac*.*2* enterotype, as well as statin use and *Bac*.*2* enterotype among obese participants, multivariable logistic regression models were generated through the statsmodels module in Python with *Bac*.*2* membership (versus all other enterotypes) as the dependent variable.

When testing for significant enterotype-by-statin interactions, HMG and metabolic parameters (blood glucose, blood insulin, HOMA-IR, and HbA1c) were log transformed prior to fitting the models. Analysis of Variance (ANOVA) or covariance (ANCOVA) models were then used to test for significant interactions (ANOVA (*measure ∼ statin_use+enterotype+statin_use*enterotype*) for unadjusted models and ANCOVA (*measure∼covariate 1+covariate 2+*… *covariateX+ statin_use+ enterotype+ statin_use*enterotype)* for covariate adjusted models)) using the statsmodels module in Python. If a significant interaction was present, post-hoc comparisons were performed between statin users and non-users within each enterotype on the covariate adjusted values (residuals) using two-sample t-tests, with Bonferroni corrected *P<0*.*05* considered statistically significant.

## Data Availability

Qualified researchers can access the full Arivale deidentified dataset supporting the findings in this study for research purposes through signing a Data Use Agreement (DUA). Inquiries to access the data can be made at and will be responded to within 7 business days.

https://github.com/gibbons-lab/mbtools

https://github.com/PriceLab/Statins_microbiome_project

## Acknowledgments

We kindly thank Arivale participants who consented to let their deidentified data be used for research purposes. This work was supported by the M.J. Murdock Charitable Trust (to L.H. and N.D.P.), and Arivale. S.M.G., and C.D. were supported by a Washington Research Foundation Distinguished Investigator Award and by start-up funds from the Institute for Systems Biology. Further support came from the National Academy of Medicine Catalyst Award (to E.S.O, S.M.G., and L.H.) and a National Institutes of Health (NIH) grant (no. U19AG023122) awarded by the National Institute on Aging (NIA) (to P.S., E.S.O, and N.R.).

## Author contributions

T.W., N.D.P., N.R., A.T.M., and S.M.G. conceptualized the study. T.W., S.K., J.C.L., P.S., E.S.O, L.H., N.D.P., N.R., A.T.M. and S.M.G. participated in the study design. T.W., S.K., C.D., M.C. performed data analysis. A.T.M., S.K., M.C., and J.C.L. managed the logistics of data collection and integration. T.W., N.R., A.T.M. and S.M.G. were the primary authors of the paper, with contributions from all others. All authors read and approved the final manuscript.

## Data and materials availability

Code used to analyze 16S rRNA gene amplicon sequencing data can be found at https://github.com/gibbons-lab/mbtools while code used to run the statistical analysis is available at https://github.com/PriceLab/Statins_microbiome_project. Qualified researchers can access the full Arivale deidentified dataset supporting the findings in this study for research purposes through signing a Data Use Agreement (DUA). Inquiries to access the data can be made at and will be responded to within 7 business days.

---

**Fig. S1.**
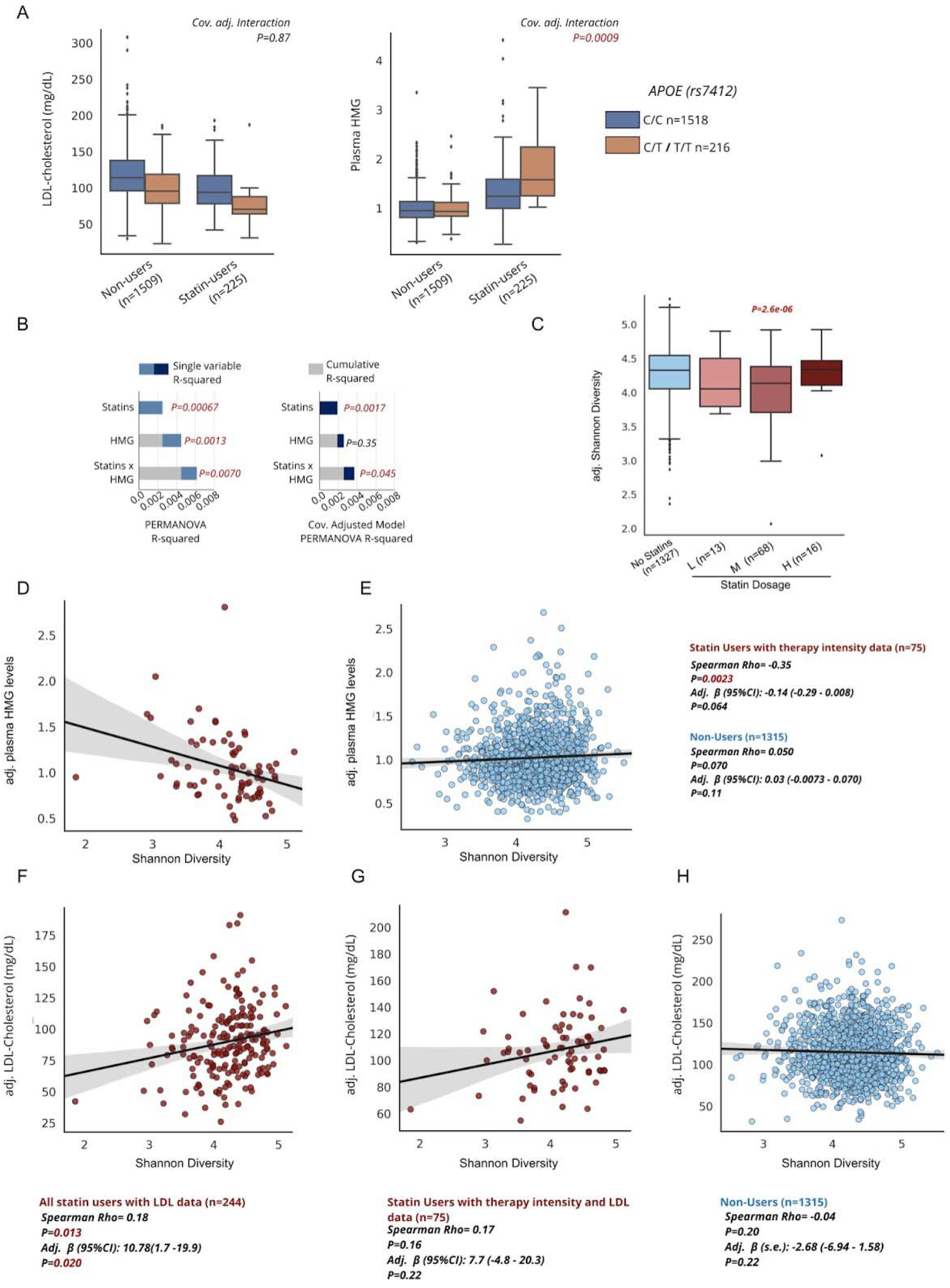
Gut alpha-diversity is anti-correlated with markers of statin on-target effects. **A)** LDL-cholesterol and plasma HMG measures in individuals stratified by statin use and genotype. Provided is the P-value for the statin-by-SNP interaction term from GLM (HMG) or OLS (LDL) models adjusted for sex, age, BMI and the first 7 ancestry principle components. **B)** Proportion of variance explained by statin use, plasma HMG levels, and a statin-by-HMG interaction term from unadjusted PERMANOVA models (statin use + HMG + statin use-by-HMG) or models adjusted for sex, age, BMI, and microbiome vendor using the Bray-Curtis genus-level dissimilarity matrix. Grey area corresponds to the cumulative R-squared of variables added to the model prior to the variable indicated on the x-axis, while the colored areas of the bars represent the additional variance explained by that variable. **C)** Measures of Observed ASVs in non-users and across statin users with known therapy intensity (low, moderate, high). **D-E)** Scatter plots of Shannon diversity (x-axis) and covariate adjusted plasma HMG levels (y-axis) in statin users with known dosage therapy intensity (D) and statin non-users (E). HMG values have been adjusted for the same covariates as in B), as well as statin intensity. Also provided are the spearman correlation coefficients and their corresponding P-value, as well as adjusted β-coefficients from GLM predicting HMG levels adjusted for the same covariates as in C) as well as dosage intensity. **F-G)** Scatter plots of Shannon diversity (x-axis) and covariate adjusted LDL-cholesterol levels (y-axis) in all statin users (F) and statin users with known therapy intensity (G), where LDL values were further adjusted for therapy intensity. **F)** Scatter plot of Shannon diversity (x-axis) and covariate adjusted LDL-cholesterol (y-axis) in statin non-users adjusted for the same covariates as in F).

**Fig. S2.**
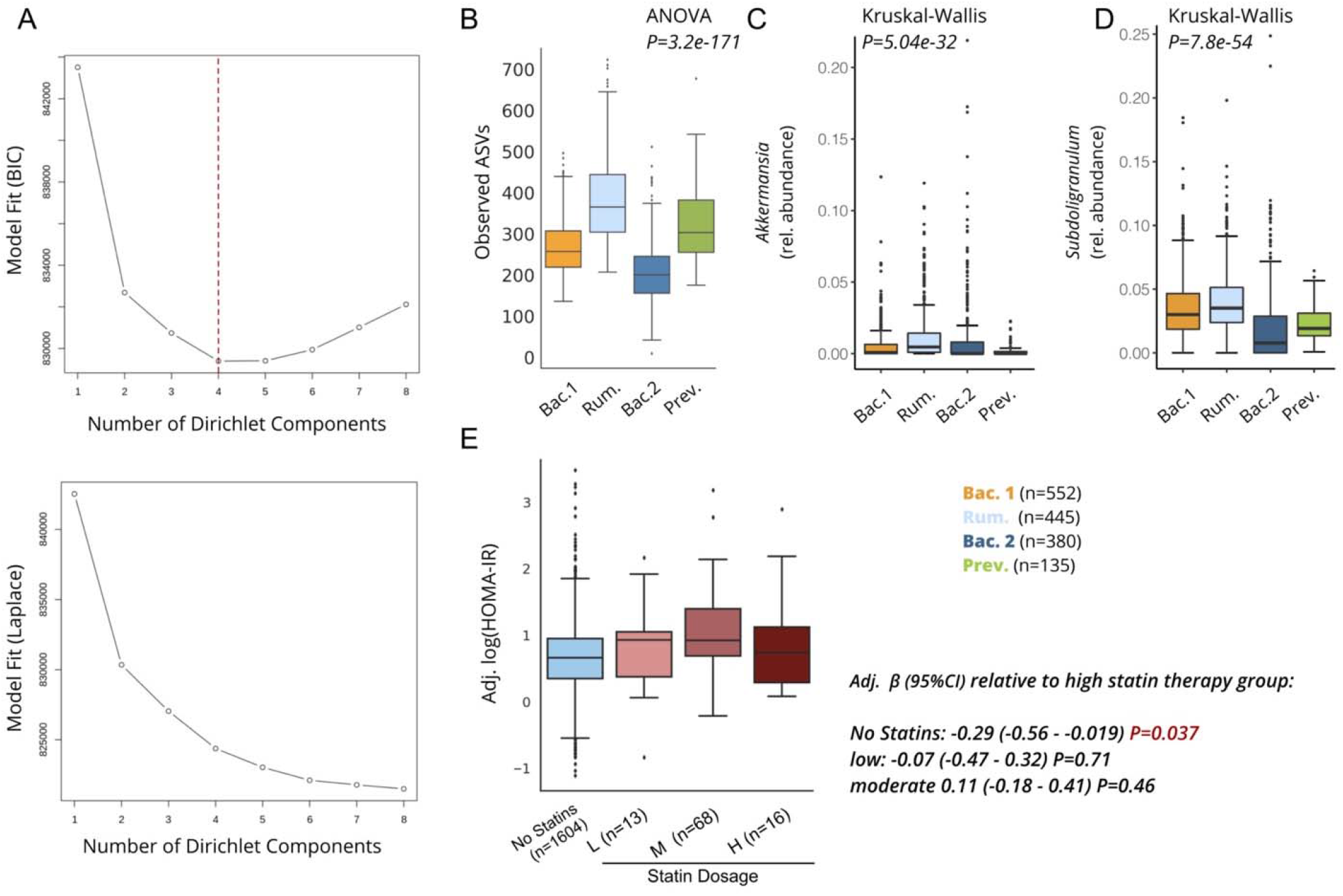
Enterotypes differ in their relative abundance of SCFA-producing taxa. **A)** Measure of model fit using the Bayesian information criterion (BIC) (top) across an increasing number of Dirichlet components as well as Laplace approximation (bottom) in the Arivale cohort. Specifying 4 components resulted in best model performance using BIC and is highlighted by the dotted red line. **B)** Gut alpha-diversity measures using Observed ASVs across the four enterotypes. **C-D)** Relative abundance of the genus *Akkermansia* (C) and *Subdoligranulum* (D) across the four enterotypes identified in the Arivale cohort. P-value from a non-parametric Kruskal-Wallis test comparing differences across all four enterotypes is provided in the top right corner. **D)** HOMA-IR levels across statin non-users and statin users with known therapy intensity. To the right are the β-coefficients, 95% confidence intervals, and P-values from OLS regression models predicting log(HOMA-IR) adjusted for clinical lab vendor, microbiome vendor, sex, age, BMI, and LDL cholesterol. HOMA-IR values on the y-axis have been adjusted for the same covariates. Box plots represent the interquartile range (25th to 75th percentile, IQR), with the middle line denoting the median; whiskers span 1.5 × IQR, points beyond this range are shown individually.

**Fig. S3:**
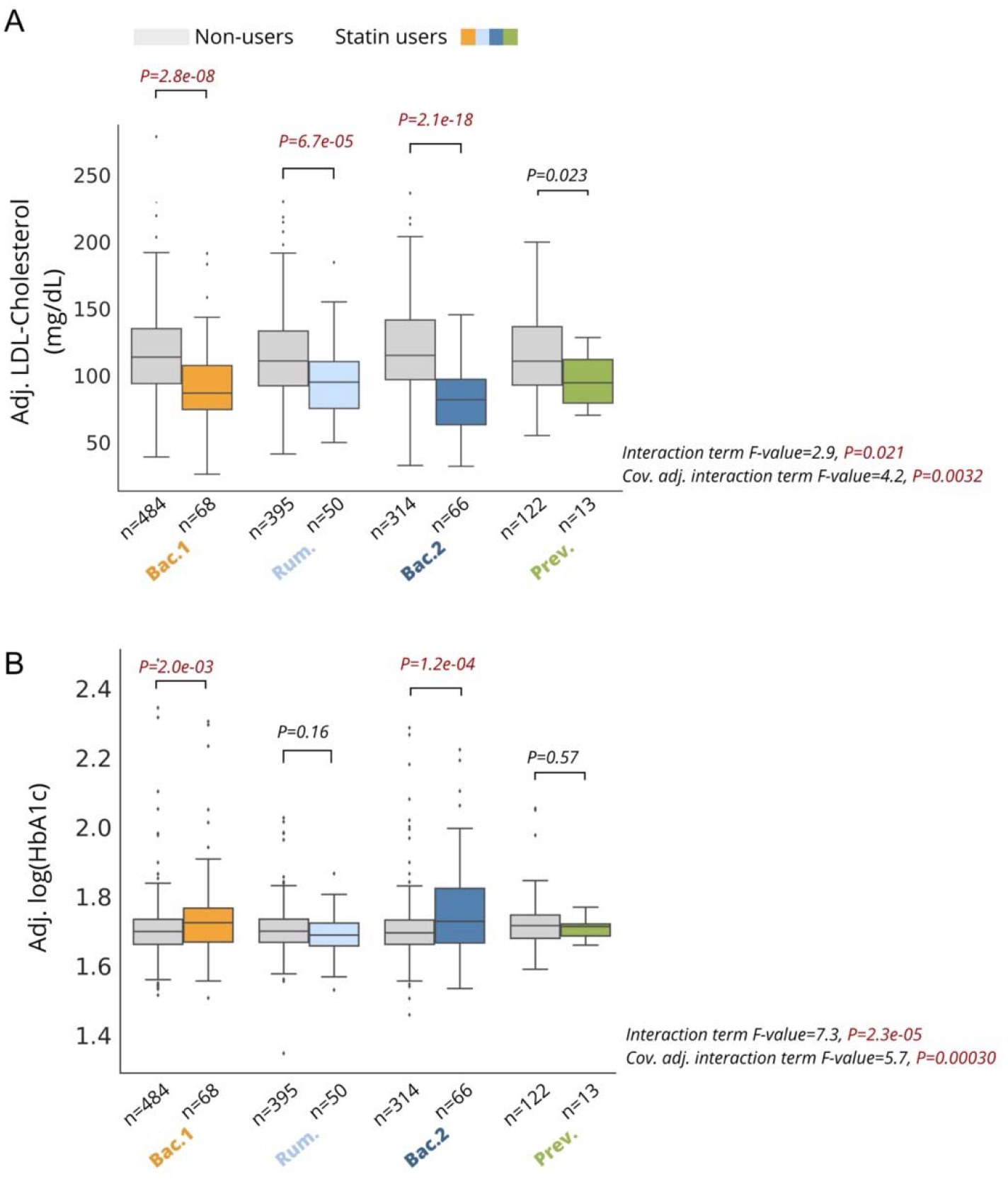
Microbiome enterotypes modify markers of statin on and off-target effects. **A)** Blood LDL-cholesterol levels among statin users and non-users stratified by enterotype. Interaction P corresponds to the statin-by-enterotype interaction term P-value from unadjusted ANOVA models, while the cov. Adj. interaction P corresponds to the statin-by-enterotype interaction term P-value from ANCOVA models adjusted for clinical lab vendor, microbiome vendor, sex, age, BMI and LDL cholesterol. Values shown on the y-axis are values adjusted for the same covariates (residuals). **B)** HbA1c measures among statin users and non-users stratified by enterotype. Interaction P corresponds to an unadjusted interaction term P-value as in A), while the cov. Adj. interaction P corresponds to the statin-by-enterotype interaction term P-value from ANCOVA models adjusted for clinical lab vendor, microbiome vendor, sex, age, BMI, HMG and LDL cholesterol. Values shown on the y-axis are values adjusted for the same covariates (residuals). P-values above the box plots across A)-B) correspond to tests of significance between statin non-users and statin users within each enterotype using two-samples t-test on covariate adjusted values (residuals). Differences with Bonferroni corrected P<0.05 were considered statistically significant and are highlighted in red. Box plots represent the interquartile range (25th to 75th percentile, IQR), with the middle line denoting the median; whiskers span 1.5 × IQR, points beyond this range are shown individually.

**Table S1.**
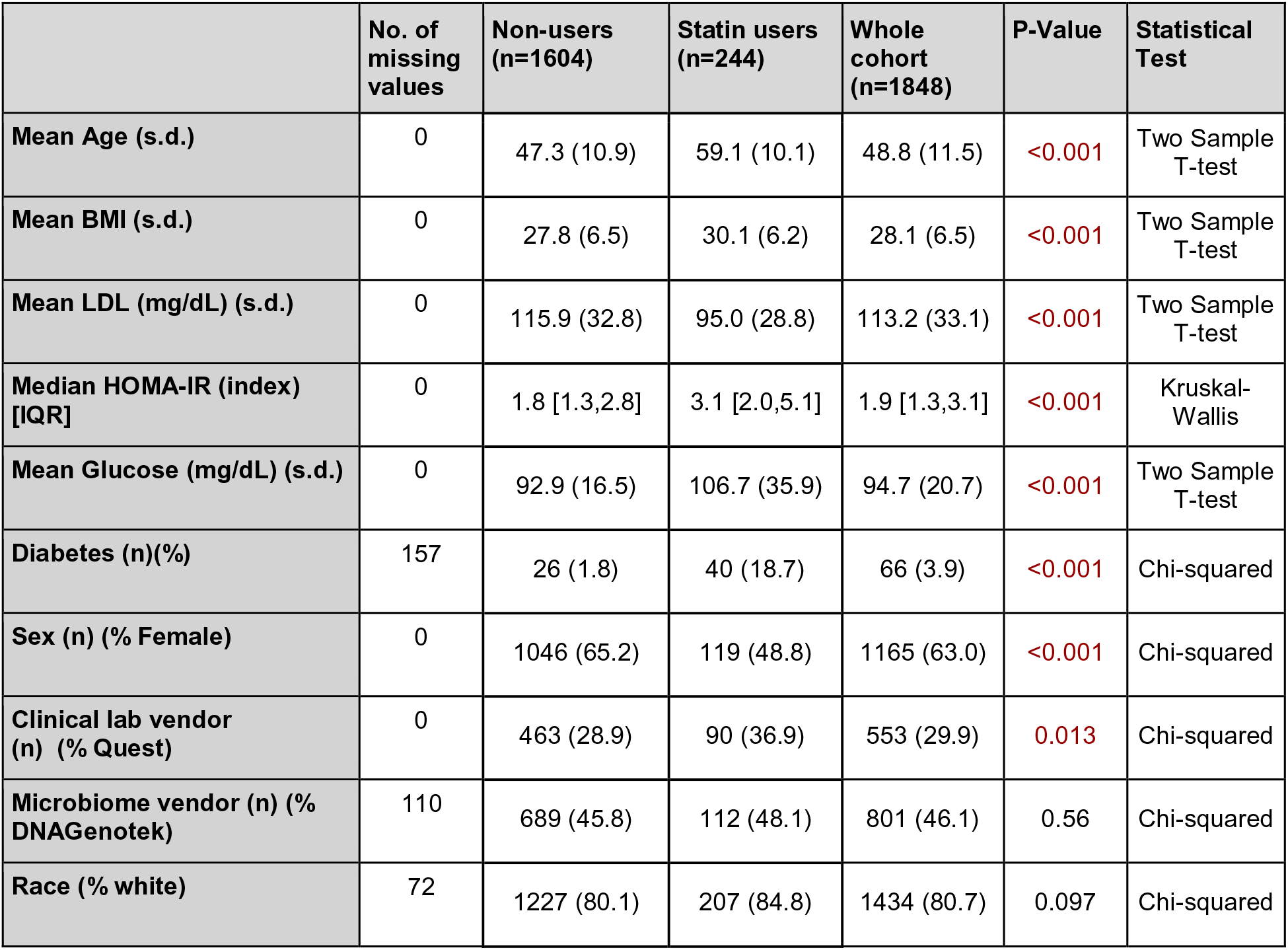
Arivale participant demographics stratified by statin use. No. of missing values corresponds to the total number of missing values across the cohort due to either participants not providing that information (diabetes status, race) or not having that omics data available (microbiome). ‘P-Value’ corresponds to statistical analysis testing the difference between statin users and non-users, with the type of statistical test used shown in the last column. Abbreviations: BMI: body mass index; LDL: low-density lipoprotein cholesterol; HOMA-IR: Homeostatic Model Assessment for Insulin Resistance; IQR: interquartile range.

|  | No. of missing values | Non-users (n=1604) | Statin users (n=244) | Whole cohort (n=1848) | P-Value | Statistical Test |
| --- | --- | --- | --- | --- | --- | --- |
| Mean Age (s.d.) | 0 | 47.3 (10.9) | 59.1 (10.1) | 48.8 (11.5) | <0.001 | Two Sample T-test |
| Mean BMI (s.d.) | 0 | 27.8 (6.5) | 30.1 (6.2) | 28.1 (6.5) | <0.001 | Two Sample T-test |
| Mean LDL (mg/dL) (s.d.) | 0 | 115.9 (32.8) | 95.0 (28.8) | 113.2 (33.1) | <0.001 | Two Sample T-test |
| Median HOMA-IR (index) [IQR] | 0 | 1.8 [1.3,2.8] | 3.1 [2.0,5.1] | 1.9 [1.3,3.1] | <0.001 | Kruskal-Wallis |
| Mean Glucose (mg/dL) (s.d.) | 0 | 92.9 (16.5) | 106.7 (35.9) | 94.7 (20.7) | <0.001 | Two Sample T-test |
| Diabetes (n)(%) | 157 | 26 (1.8) | 40 (18.7) | 66 (3.9) | <0.001 | Chi-squared |
| Sex (n) (% Female) | 0 | 1046 (65.2) | 119 (48.8) | 1165 (63.0) | <0.001 | Chi-squared |
| Clinical lab vendor (n) (% Quest) | 0 | 463 (28.9) | 90 (36.9) | 553 (29.9) | 0.013 | Chi-squared |
| Microbiome vendor (n) (% DNAGenotek) | 110 | 689 (45.8) | 112 (48.1) | 801 (46.1) | 0.56 | Chi-squared |
| Race (% white) | 72 | 1227 (80.1) | 207 (84.8) | 1434 (80.7) | 0.097 | Chi-squared |

**Table S2.** Correspondence of HMG with SNPs previously associated with statin response. β-coefficients, standard error (s.e.) and the corresponding p-value for the SNP-by-statin interaction term predicting either HMG (GLM) or LDL-cholesterol levels (OLS regression) across the Arivale cohort with available genetics data. Models were adjusted for sex, age, BMI and the first 7 ancestry PCs. “Corr. P-value” corresponds to the P-value for each β-coefficient after correcting for multiple hypothesis testing (FDR<0.05). Significant P-values are highlighted in red.

|  |  | HMG (N=1734) |  |  |  | LDL-Cholesterol (N=1734) |  |  |  |
| --- | --- | --- | --- | --- | --- | --- | --- | --- | --- |
| SNP rsid | At least one copy of the minor allele (proportion) | Adj. β-coeff | s.e. | P-value | Corr. P-value | Adj. β-coeff | s.e. | P-value | Corr. P-value |
| rs10455872 | 0.11 | -0.0207 | 0.0338 | 0.5402 | 0.6690 | -4.5819 | 3.5322 | 0.1947 | 0.4333 |
| rs2199936 | 0.23 | -0.0157 | 0.0245 | 0.5211 | 0.6690 | -0.2432 | 2.5747 | 0.9247 | 0.9966 |
| rs2900478 | 0.29 | -0.0110 | 0.0237 | 0.6427 | 0.6690 | 1.0850 | 2.4733 | 0.6609 | 0.9966 |
| rs4420638 | 0.30 | -0.0178 | 0.0228 | 0.4350 | 0.6690 | -3.4403 | 2.3689 | 0.1466 | 0.4333 |
| rs445925 | 0.18 | 0.0907 | 0.0324 | 0.0052 | **0.0207** | -0.0143 | 3.3645 | 0.9966 | 0.9966 |
| rs7412 | 0.12 | 0.1513 | 0.0453 | 0.0009 | **0.0068** | 0.7567 | 4.6668 | 0.8712 | 0.9966 |
| rs646776 | 0.37 | 0.0096 | 0.0224 | 0.6690 | 0.6690 | 4.1731 | 2.3359 | 0.0742 | 0.4333 |
| rs8014194 | 0.46 | 0.0347 | 0.0215 | 0.1068 | 0.2849 | 2.7833 | 2.2520 | 0.2166 | 0.4333 |

**Data S1. Significant genus differences across enterotypes**. List of 85 genera significantly differing across enterotypes tested using a Kruskal-Wallis test (Bonferroni *P<0*.*05*). Each enterotype column corresponds to the median relative abundance of a particular genus for that enterotype.

